# The Role and Regulation of Intramuscular Sex Hormones in Skeletal Muscle: A Systematic Review

**DOI:** 10.1101/2024.12.07.24318664

**Authors:** Viktor Engman, Annabel J. Critchlow, Eija K. Laakkonen, Mette Hansen, Shaun Mason, Séverine Lamon

## Abstract

**Introduction:** Serum concentrations of androgens and oestrogens, the main male and female sex hormones, respectively, naturally fluctuate across the lifespan. Sex hormones are mainly produced in the gonads, but evidence suggests that they can also be locally synthesised in skeletal muscle. However, little is known about the purpose of intramuscular sex hormones biosynthesis and their role in skeletal muscle. This systematic review aimed to investigate 1) how intramuscular sex hormone levels vary across the lifespan, 2) whether intramuscular sex hormones are associated with skeletal muscle mass and function, and 3) whether exercise affects intramuscular sex hormone levels.

**Methods:** Four databases were searched, and studies were included if they contained measurements of intramuscular sex hormones from healthy males and females free from any hormonal treatment, from rodents, or from cultured muscle cells.

**Results:** Fifteen studies were included. Intramuscular testosterone was reduced in elderly males compared to their younger counterparts, but comparison of intramuscular sex hormone levels between pre- and postmenopausal females yielded inconclusive findings. Intramuscular androgens were positively associated with muscle mass and strength in males. In females, conflicting findings were reported for both oestradiol and androgens, and measures of muscle mass and function. Chronic exercise decreased androgens and oestradiol in females, but increased androgens in males. Acute exercise did not change intramuscular hormone levels in humans but increased them in rodents and cells.

**Conclusion:** Current evidence suggests that ageing and exercise differentially modulate intramuscular sex hormone levels, and their association with muscle mass and function, between males and females.

## 2 Introduction

Maintenance of muscle mass and function is an integral part of healthy ageing. Across adulthood, muscle is lost at a median decennial rate of 4.7% in males and 3.7% in females,^1^ which is associated with increased mortality,^2^ loss of independence and reduced quality of life.^3,4^ In comparison to younger adults, older individuals exhibit a diminished anabolic response to hypertrophic stimuli, such as resistance exercise and protein intake,^5,6^ which impairs their capacity to counteract age-related muscle loss.

Skeletal muscle is a particularly sex-biased organ with nearly 3000 genes differentially expressed between males and females in a postabsorptive state.^7^ Consequently, males and females present distinct muscle phenotypes, which differ in terms of fibre type proportions, substrate metabolism, contractile speed and fatigability.^8–10^ The rate of age-related muscle mass and functional decline is also sex-dependent,^11,12^ with atrophy and functional impairments being more pronounced and severe in postmenopausal females.^13^ Even though the ageing skeletal muscle transcriptomic signature is largely similar between sexes, the extent of the differentially expressed genes between young and older adults is sex-dependent.^14,15^

Physiological sex differences emerge as early as the first trimester of gestation but are accentuated during puberty, concomitant to the elevated secretion of sex steroid hormones,^16–20^ predominantly androgens and oestrogens. After puberty, the serum levels of the androgen hormone testosterone is ∼15 times higher in males than females.^21^ Endogenous serum testosterone levels are positively associated with muscle mass and strength in males,^22,23^ but not in females.^24^ However, supraphysiological testosterone levels achieved through exogenous administration have significant anabolic effects in both sexes, resulting in substantial and rapid increases in muscle mass and strength.^25–28^

The most potent of oestrogens, oestradiol (E2), fluctuates in the blood during the menstrual cycle in premenopausal females and is markedly enhanced during pregnancy.^29^ Otherwise, E2 remains relatively stable during the reproductive years of adulthood.^30,31^ During menopause, females experience a rapid and permanent decline in E2 levels whereas male levels are unaltered with ageing.^31^ The relationship between serum oestrogen and indices of muscle mass and function remains equivocal, with conflicting findings regarding both muscle mass and strength in females,^32^ and no apparent association in males.^33,34^

Serum sex hormone levels may be modified through exercise. Initiation of resistance training may increase basal testosterone levels in males but not females,^35,36^ which may progressively increase for years with intense exercise training.^36,37^ Even though basal testosterone levels are associated with muscle hypertrophy and strength gains,^25,28^ muscle hypertrophy can occur when endogenous testosterone levels are suppressed, albeit to a lesser magnitude.^38^ Others have suggested that the exercise induced elevations in androgen receptor (AR) content might be a determining factor for hypertrophic potential.^39,40^ More recently, nuclear AR content was found to correlate with muscle hypertrophy in males, but not females, highlighting a potential sex-based discrepancy in the mechanisms of muscle hypertrophy.^41,42^

Serum E2 levels acutely increase during endurance exercise in both young females and males.^43^ In females, the increments appear to be mediated by the menstrual status. Nakamura *et al.*^44^ found that in eumenorrheic females, acute increments in circulating E2 levels were only detectable in the mid-luteal phase and absent in the early follicular phase, while E2 remains unchanged following acute exercise in postmenopausal females.^45^ Further, E2 was also unchanged in amenorrhoeic females following acute exercise.^44^ The acute exercise-induced increases in E2 observed in eumenorrheic females and males may be transient, as longitudinal training studies have failed to replicate such results over time.^46,47^

Classically, skeletal muscle was thought of as merely a target tissue for sex hormone signalling, as it expresses both the AR and oestrogen receptors (ERs).^39,48,49^ However, extragonadal biosynthesis of steroid hormones occurs in a plethora of tissues,^50^ including skeletal muscle. Aizawa *et al.* (2007)^51^ first showed that skeletal muscle from male rats expresses the necessary enzymes, namely 3-beta-hydroxysteroid dehydrogenase (3β-HSD) and 17-beta-hydroxysteroid dehydrogenase (17β-HSD), to synthesise testosterone from its precursor dehydroepiandrosterone (DHEA) and aromatase cytochrome P-450 (P450arom) to convert testosterone into E2. Using DHEA treated rat myotubes, they also demonstrated a dose response relationship between DHEA concentration and subsequent intramuscular levels of testosterone and E2. Further, 3-oxo-5α-steroid 4-dehydrogenase (5α-reductase), the enzyme enabling the conversion of testosterone to the more potent DHT, is also expressed in cultured rodent skeletal muscle.^52^ The expression of these enzymes has subsequently been confirmed in human male and female skeletal muscle.^53,54^

The role, regulation, and function of intramuscular sex hormones on skeletal muscle health is yet to be systematically reviewed to further our understanding of how sex hormones might relate to muscle mass and function over the lifespan. The objectives of this review were threefold and aimed to address the following research questions: 1) whether intramuscular sex hormones vary across the life span in humans; 2) whether intramuscular sex hormones are associated with muscle mass and function or increments in these, and 3) whether exercise impacts intramuscular sex hormone levels.

## 3 Methods

### 3.1 Search Strategy

The systematic review was registered at PROSPERO (CRD42023479951). Four databases (MEDLINE Complete, EMBASE, SPORTDiscus, and PubMed) were searched according to the full search criteria outlined in supplementary table S1. The databases were searched until October 27^th^, 2023.

#### 3.1.1 Inclusion and exclusion criteria

In brief, study subjects included healthy adults, rodent models and *in vitro* cell models all free from hormonal treatments. Measures of intramuscular sex hormones and/or enzymes involved in the hormone biosynthesis were required for all included studies. Different eligibility criteria were applied to answer the different research questions as outlined in supplementary table S2.

#### 3.1.2 Screening and selection

Following the database search, the relevant papers were imported into Covidence systematic review software (Veritas Health Innovation, Melbourne, Australia).^55^ Duplicates were automatically removed. Two authors independently assessed eligibility through title and abstract screening and subsequently full-text assessment and removed irrelevant studies according to the pre-specified eligibility criteria. The PRISMA search diagram has been outlined in figure 1.

**Figure 1.**
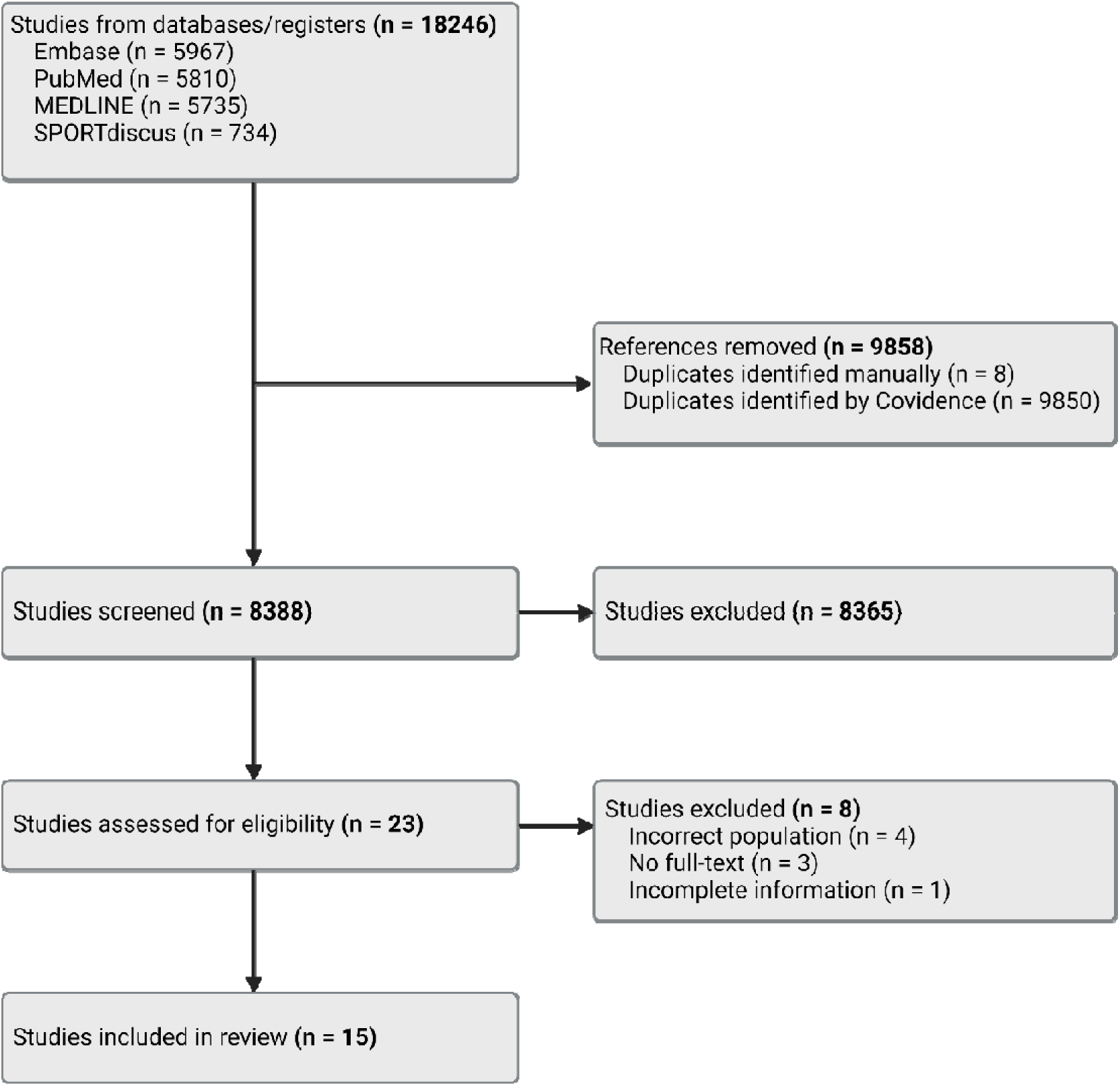
PRISMA diagram outlining the systematic review search and assessment.

#### 3.1.3 Data extraction and quality assessment

Information about study design, study subjects, methodology, and results were independently extracted by two authors from all eligible studies using a predesigned data extraction form in Covidence. Any conflicts between the two authors were resolved by jointly extracting the data from the relevant study.

Quality assessment was conducted using the appropriate Joanna Briggs Institute (JBI) critical appraisal tool for human studies based on the study design (cross sectional, randomised controlled trial, quasi experimental trial).^56,57^ Here, the percentage of “yes” answers were tallied and scored accordingly: ≤49% yes = high risk of bias, 50-69% yes = moderate risk of bias, ≥70% yes = low risk of bias. Animal studies were appraised using SYRCLE’s risk of bias tool for animal studies and *in vitro* studies using an adapted version of the JBI Randomised Controlled Trial critical appraisal tool.^58^

#### 3.1.4 Methods of synthesis

During the data extraction process, the included studies were assigned, analysed, and discussed according to each of the relevant aims. Due to anticipated study heterogeneity, study outcomes were not combined quantitively into a meta-analysis. The strength of associations between intramuscular hormones and measures of muscle mass and function was converted to Pearson’s correlation coefficient where applicable and referred to as small (r≥0.1 and <0.3), moderate (r≥0.3 and <0.5), or large (r≥0.5).

## 4 Results

### 4.1 Study characteristics

Following the database search and screening, 23 studies were included for full-text analysis, and 15 studies were deemed eligible. Three studies investigated how intramuscular sex hormones varied across the lifespan (aim 1),^53,54,59^ eight studies investigated the putative relationship between intramuscular hormones and muscle mass and function (aim 2),^39,45,53,54,59–62^ and 13 studies investigated the relationship between exercise and intramuscular sex hormone concentrations (aim 3).^39,45,54,60–69^ Eight studies were excluded due to unavailability of a full-texts (n=3), incomplete information (n=1), and incorrect population (n=4).

### 4.2 Quality assessment

Using the JBI risk of bias tools, the cross-sectional human studies presented a moderate risk of bias,^53,59^ and the quasi-experimental human trials all presented a low risk of bias.^54,65–68^ Out of the two randomised controlled trials, one presented a low risk of bias whereas the other presented a high risk of bias.^39,45^ Animal trials were critically appraised using the SYRCLE tool, where all included studies had a similar risk of bias, notably missing information on researcher blinding.^60,62–64^ The two *in vitro* cell culture studies were critically appraised using a JBI adapted critical appraisal tool with variations in methodological explication.^61,69^ The full quality assessment has been outlined in supplementary tables S3-S7.

### 4.3 Intramuscular sex hormone levels across the lifespan

Three human studies provided cross sectional data and measured intramuscular sex hormones via enzyme-linked immunosorbent assay (ELISA) or enzyme immunoassays (EIA). Two of these studies examined differences in intramuscular sex hormone levels between pre- and postmenopausal females,^53,59^ and one between young and older males (Table 1).^54^ The findings were inconsistent across studies. In females, Pöllänen *et al.* (2015)^59^ found that intramuscular E2 and testosterone were similar between pre- and postmenopausal females whereas intramuscular DHT was higher in the postmenopausal females. The same group (2011)^53^ had however reported the opposite in an earlier study, where levels of intramuscular E2 and testosterone were elevated in the postmenopausal females compared to premenopausal females, and intramuscular DHT remained similar between the groups. Intramuscular DHEA did not differ between pre- and postmenopausal females in both studies.^53,59^ Compared to premenopausal females, postmenopausal females had increased intramuscular 3β-HSD mRNA levels whereas intramuscular 17β-HSD, and 5α-reductase type 1 and 2 mRNA were unaltered by menopausal status.^53^ In males, intramuscular free testosterone, DHT, DHEA, 3β-HSD, 17β-HSD, and 5α-reductase were all decreased in the older when compared to the young group.^54^

**Table 1.**
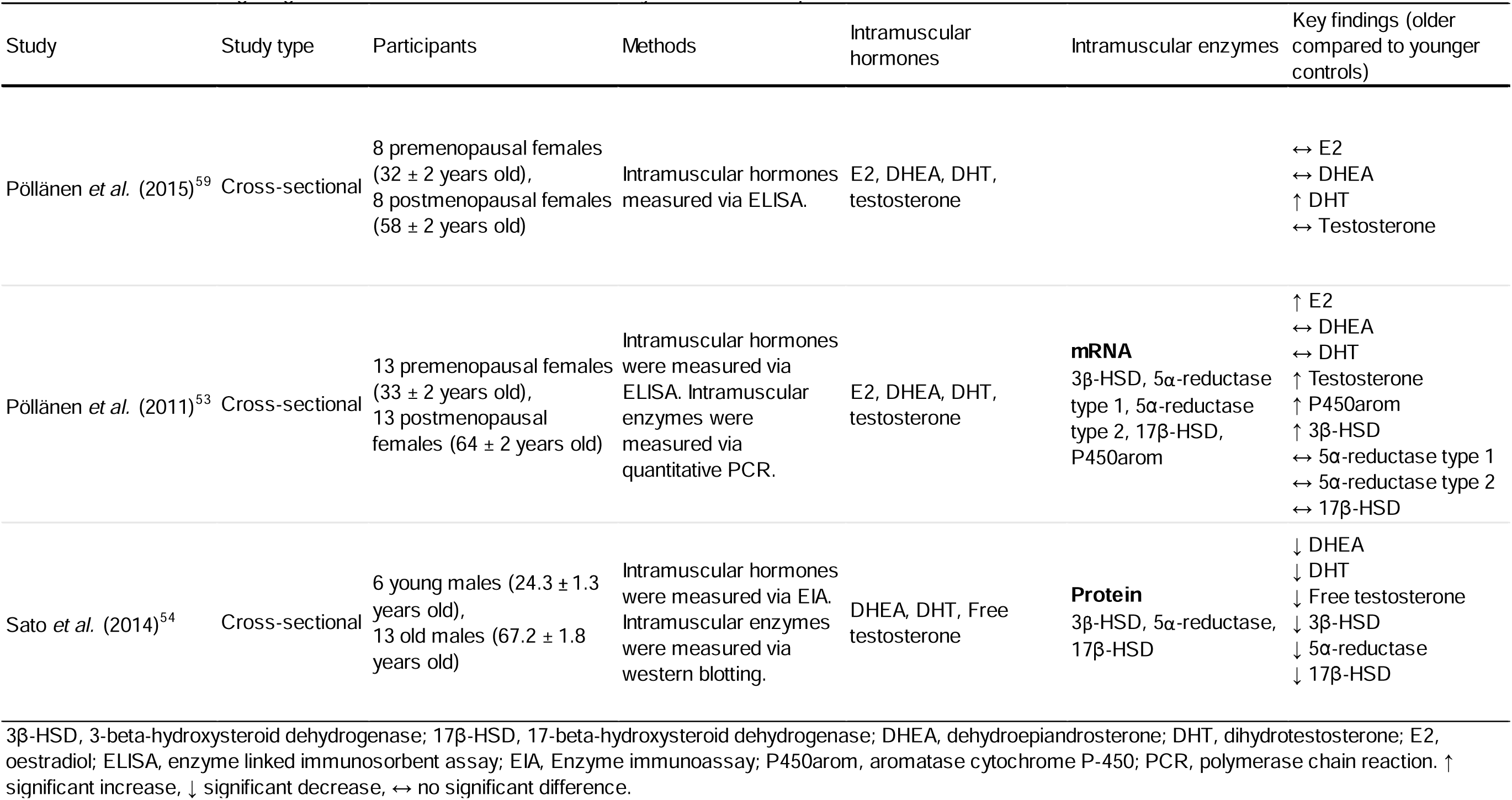
Studies investigating how intramuscular sex hormones vary across the lifespan.

| Study | Study type | Participants | Methods | Intramuscular hormones | Intramuscular enzymes | Key findings (older compared to younger controls) |
| --- | --- | --- | --- | --- | --- | --- |
| Pöllänen *et al.* (2015)<sup>59</sup> | Cross-sectional | 8 premenopausal females (32 ± 2 years old),<br>8 postmenopausal females (58 ± 2 years old) | Intramuscular hormones measured via ELISA. | E2, DHEA, DHT, testosterone |  | ↔ E2<br>↔ DHEA<br>↑ DHT<br>↔ Testosterone |
| Pöllänen *et al.* (2011)<sup>53</sup> | Cross-sectional | 13 premenopausal females (33 ± 2 years old),<br>13 postmenopausal females (64 ± 2 years old) | Intramuscular hormones were measured via ELISA. Intramuscular enzymes were measured via quantitative PCR. | E2, DHEA, DHT, testosterone | **mRNA**<br>3β-HSD, 5α-reductase type 1, 5α-reductase type 2, 17β-HSD, P450arom | ↑ E2<br>↔ DHEA<br>↔ DHT<br>↑ Testosterone<br>↑ P450arom<br>↑ 3β-HSD<br>↔ 5α-reductase type 1<br>↔ 5α-reductase type 2<br>↔ 17β-HSD |
| Sato *et al.* (2014)<sup>54</sup> | Cross-sectional | 6 young males (24.3 ± 1.3 years old),<br>13 old males (67.2 ± 1.8 years old) | Intramuscular hormones were measured via EIA. Intramuscular enzymes were measured via western blotting. | DHEA, DHT, Free testosterone | **Protein**<br>3β-HSD, 5α-reductase, 17β-HSD | ↓ DHEA<br>↓ DHT<br>↓ Free testosterone<br>↓ 3β-HSD<br>↓ 5α-reductase<br>↓ 17β-HSD |

3β-HSD, 3-beta-hydroxysteroid dehydrogenase; 17β-HSD, 17-beta-hydroxysteroid dehydrogenase; DHEA, dehydroepiandrosterone; DHT, dihydrotestosterone; E2, oestradiol; ELISA, enzyme linked immunosorbent assay; EIA, Enzyme immunoassay; P450arom, aromatase cytochrome P-450; PCR, polymerase chain reaction. ↑ significant increase, ↓ significant decrease, ↔ no significant difference.

### 4.4 Intramuscular sex hormones and muscle mass and function

Eight studies investigated the putative association between intramuscular sex hormones and muscle mass and function (Table 2).

**Table 2.**
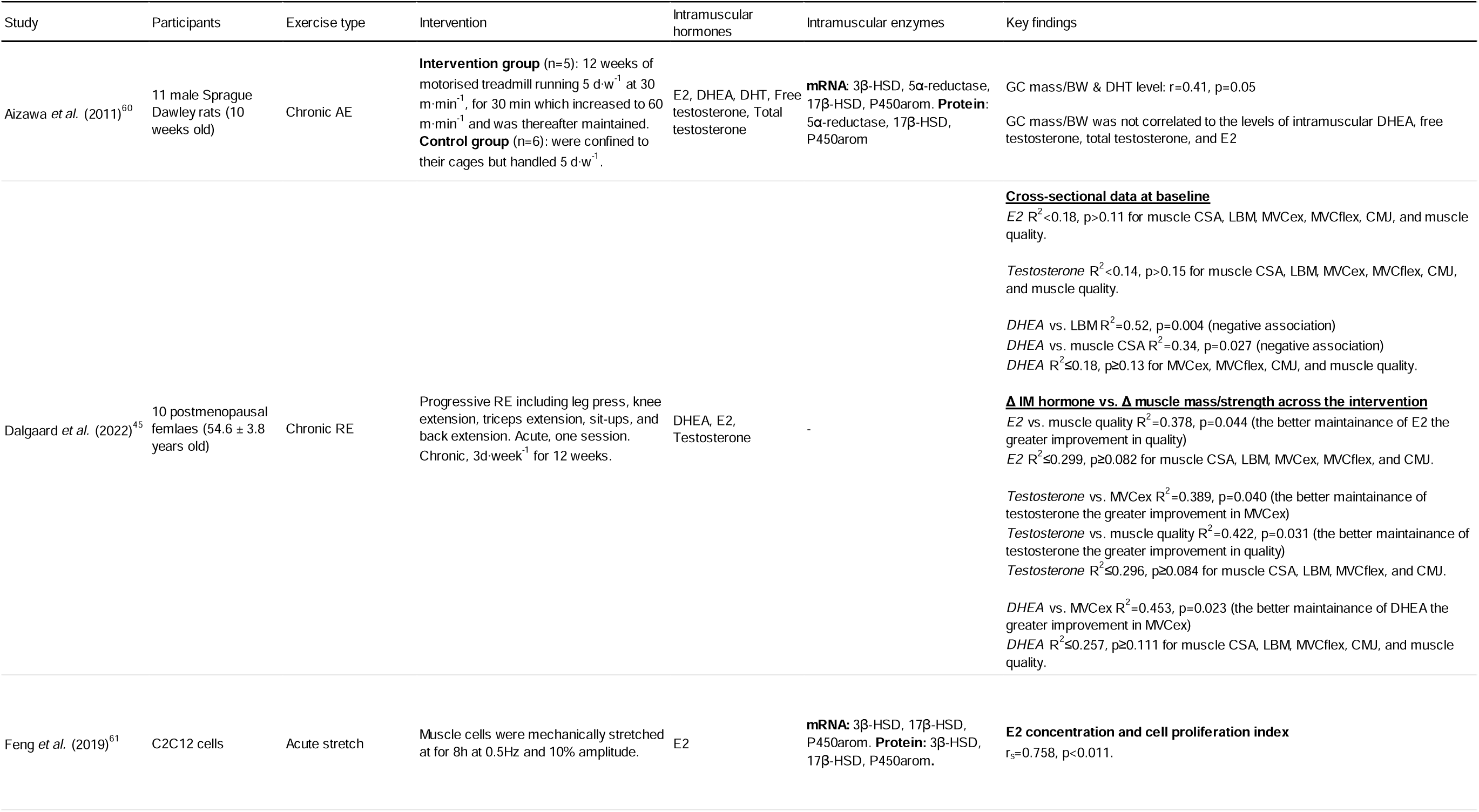
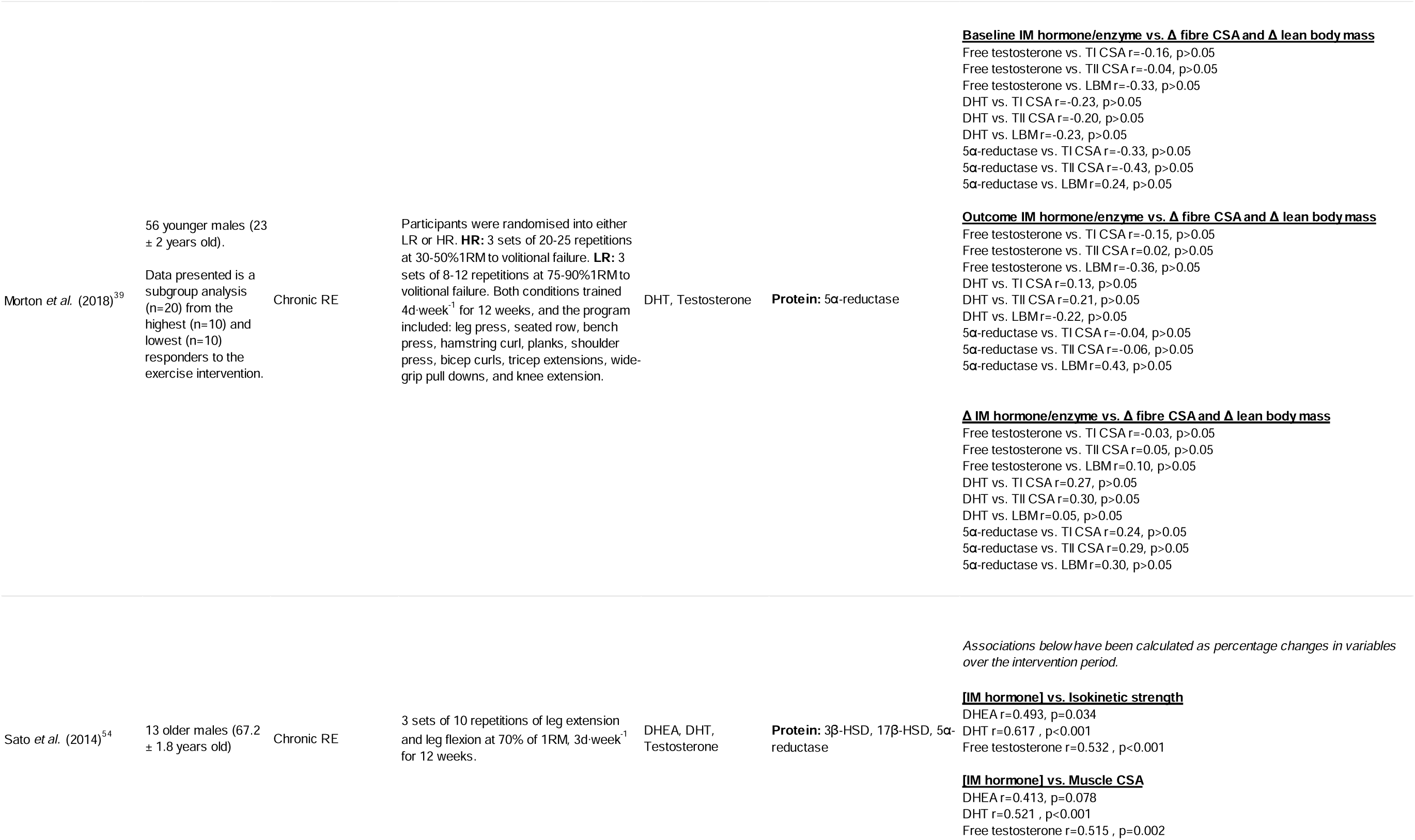
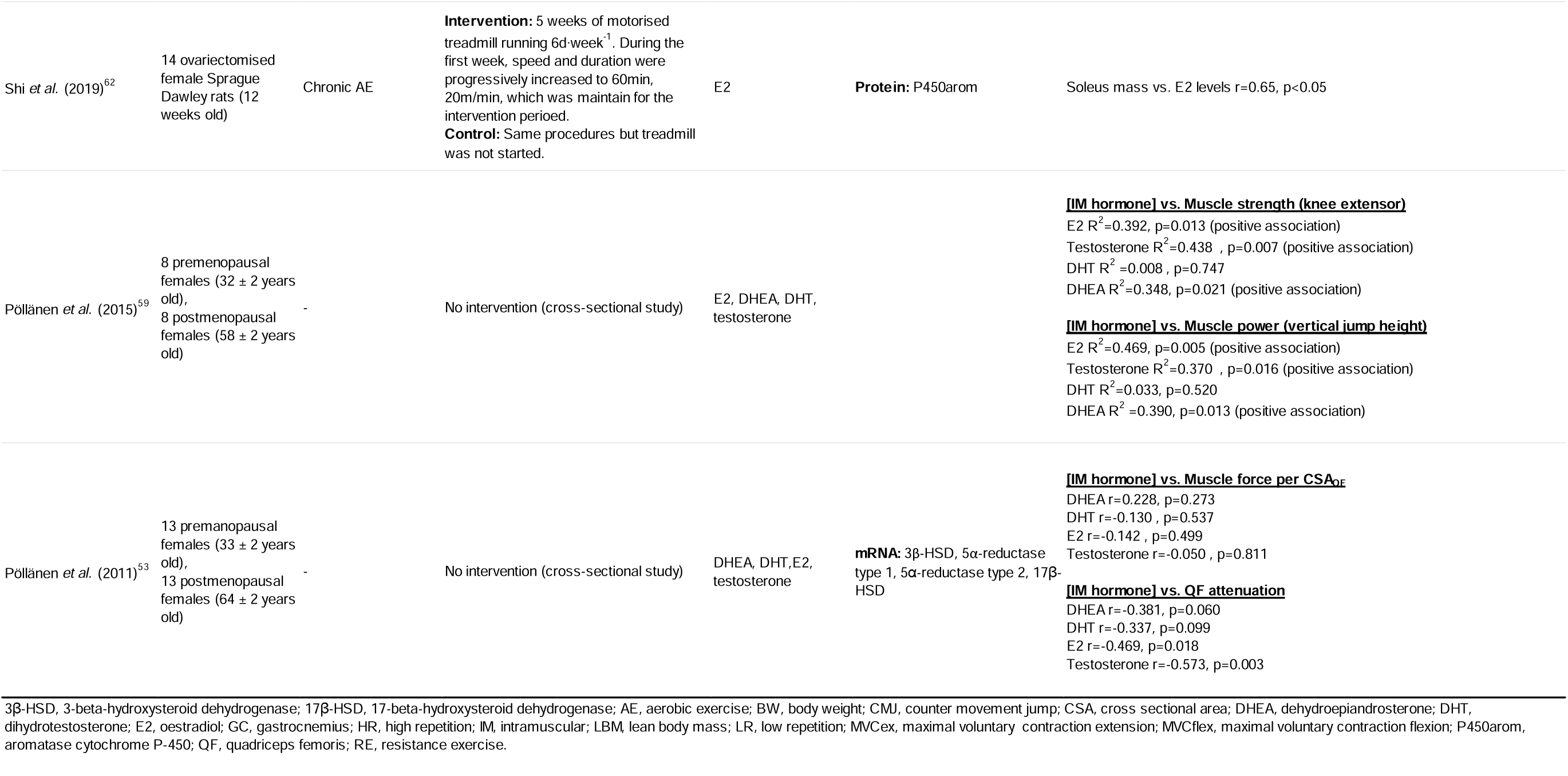
Studies investigating the relationship between intramuscular hormones and muscle mass and function.

#### 4.4.1 Muscle mass and composition

Four human studies, two animal studies, and one cell culture study assessed the putative associations between intramuscular sex hormones and measures of muscle mass and composition (Table 2). Following 12 weeks of aerobic exercise in male rats, a moderate significant positive correlation was found between gastrocnemius mass and intramuscular DHT levels.^60^ In ovariectomised female rats following 5 weeks of aerobic exercise, soleus mass presented a strong significant positive association with intramuscular E2 levels.^62^

In postmenopausal females, Dalgaard *et al.*^45^ reported large significant negative associations between intramuscular DHEA and lean body mass (LBM), and intramuscular DHEA and muscle cross sectional area (CSA), but found no such associations for intramuscular testosterone or E2. The same study found no associations between change in intramuscular DHEA, testosterone, and E2, and change in LBM and muscle CSA following a resistance training intervention.^45^ Pöllänen *et al.* (2011)^53^ assessed computed tomography attenuation (to infer fat infiltration, where a lower attenuation suggests a greater fat infiltration) in the quadriceps femoris (QF) muscle. They reported significant low to moderate negative associations between intramuscular E2 and testosterone, and QF attenuation, with no significant findings for intramuscular DHEA and DHT.^53^

In younger males, Morton *et al.*^39^ pooled data from the highest (n=10) and lowest (n=10) exercise responders and found no significant associations between baseline, post intervention, or change in intramuscular free testosterone, DHT, and 5α-reductase protein levels, and exercise induced changes in type I and II muscle fibre CSA and LBM. In older males, Sato *et al.*^54^ reported moderate to strong correlations between increments in intramuscular free testosterone and DHT, and gains in quadriceps CSA. Feng *et al.*^61^ found a strong significant positive correlation between intracellular E2 concentration and cell proliferation index in C2C12 cells following 8 hours of mechanical stretch.

#### 4.4.2 Muscle strength and function

Four human studies assessed the putative association between intramuscular sex hormones and measures of muscle strength and function (Table 2). Following a 12-week resistance training intervention in older males, the increase in intramuscular free testosterone, DHT, and DHEA all presented a significant moderate to strong positive correlation with exercise induced increments in isokinetic leg extensor torque.^54^

In postmenopausal females, Dalgaard *et al.*^45^ found no significant associations between intramuscular DHEA, testosterone, and E2, and maximal voluntary contraction (MVC) during leg flexion (MVCflex) and leg extension (MVCex), and muscle strength normalised to muscle CSA at baseline. Following an exercise intervention, maintenance of the intramuscular DHEA levels was strongly associated with improvements in MVCex, but not changes in MVCflex. Similarly, maintenance of intramuscular testosterone was strongly associated with improvements in MVCex and normalised muscle strength, but not MVCflex. Also, maintenance of intramuscular E2 was associated with improvements in normalised muscle strength.^45^

In grouped samples of pre- and postmenopausal females, intramuscular E2, testosterone and DHEA, but not DHT, presented significant strong associations with knee extensor strength.^59^ However, in another study from the same group using a similar sample and design, they reported no significant associations between intramuscular E2, testosterone, DHEA, or DHT, and knee extensor strength when normalised to quadriceps femoris CSA.^53^

Muscle power was indirectly tested by two studies. Pöllänen *et al.* (2015)^59^ found significant strong correlations between intramuscular E2, testosterone, and DHEA, but not DHT, and vertical jump height in their mixed cohort of pre- and postmenopausal females. Dalgaard *et al.*^45^ found no significant associations between any of the intramuscular DHEA, testosterone, and E2 and vertical jump height, neither at baseline nor when comparing changes in the intramuscular hormones to changes in vertical jump height, only including postmenopausal females.

### 4.5 Exercise, intramuscular sex hormones and expression of the local intramuscular sex hormone biosynthesis machinery

#### 4.5.1 Acute exercise

Thirteen studies investigated the effects of acute (n=9) and chronic (n=5) effects of exercise on intramuscular sex hormone levels in humans (n=7), rats (n=4) or cells (n=2) (Table 3). In human males and females, a single session of resistance training (n=5 studies) did not change the intramuscular levels of testosterone, E2, DHT, DHEA nor protein levels for the key enzymes 3β-HSD, 17β-HSD, P450arom and 5α-reductase.^45,65–68^

**Table 3.**
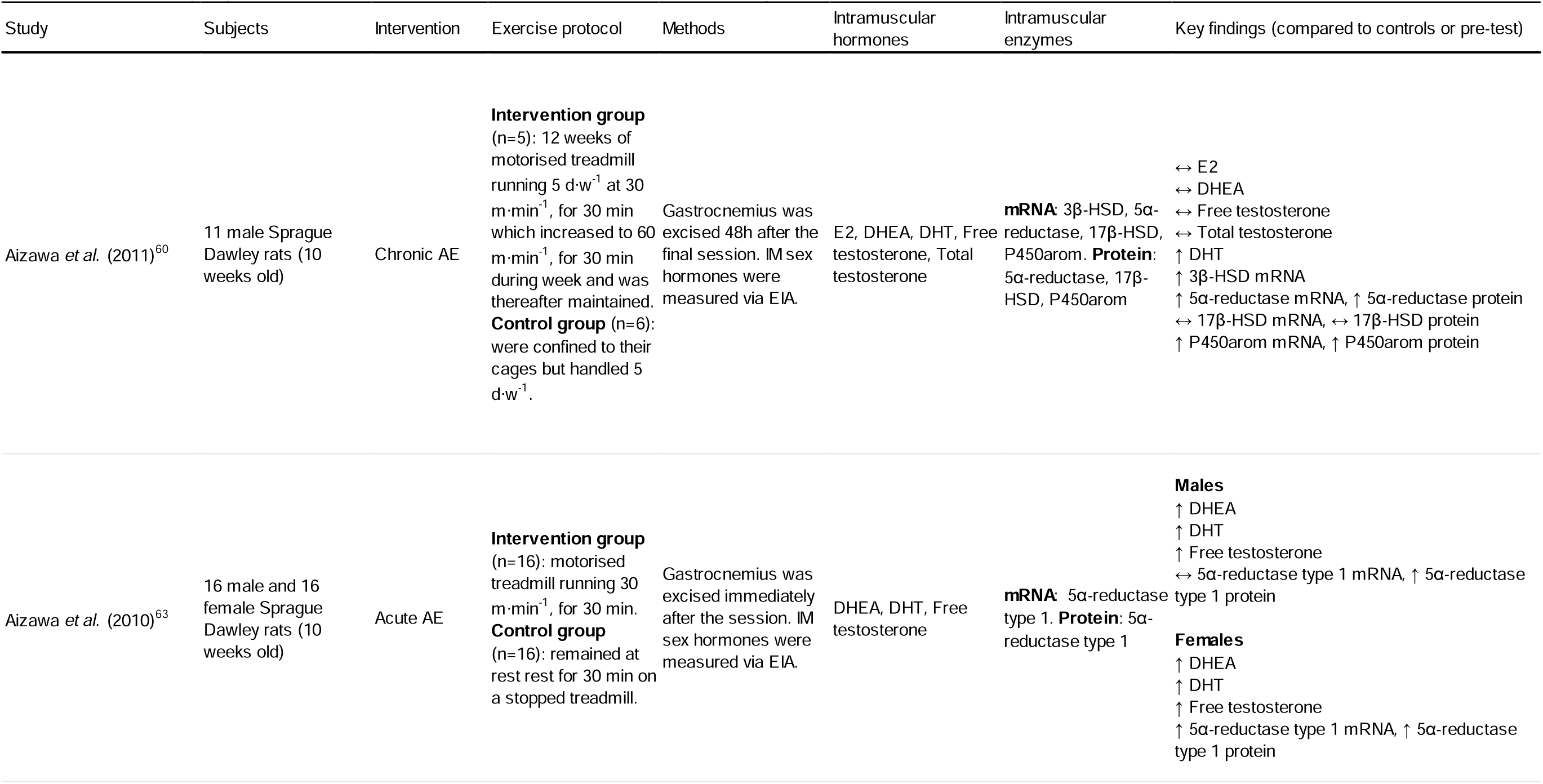
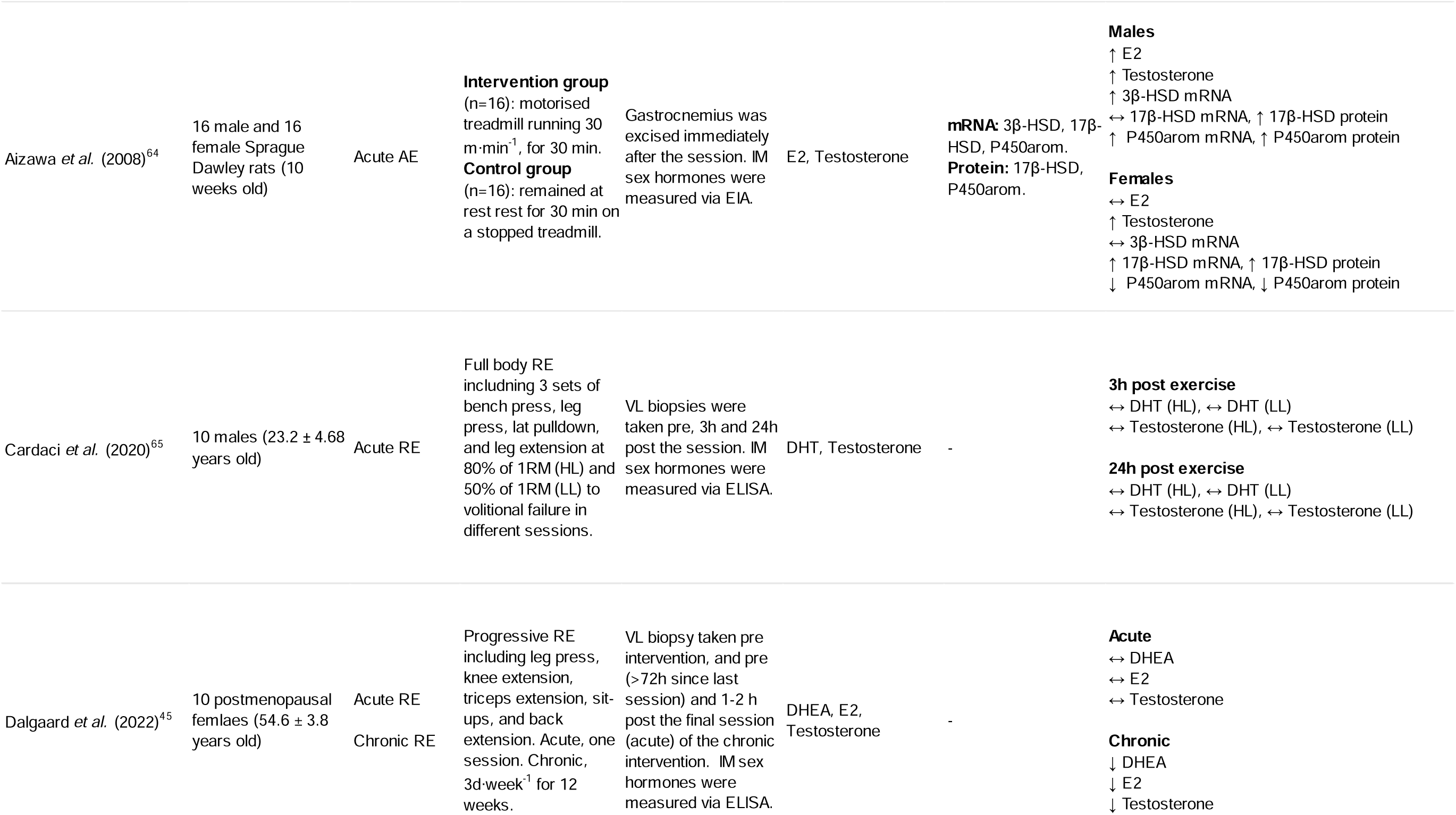
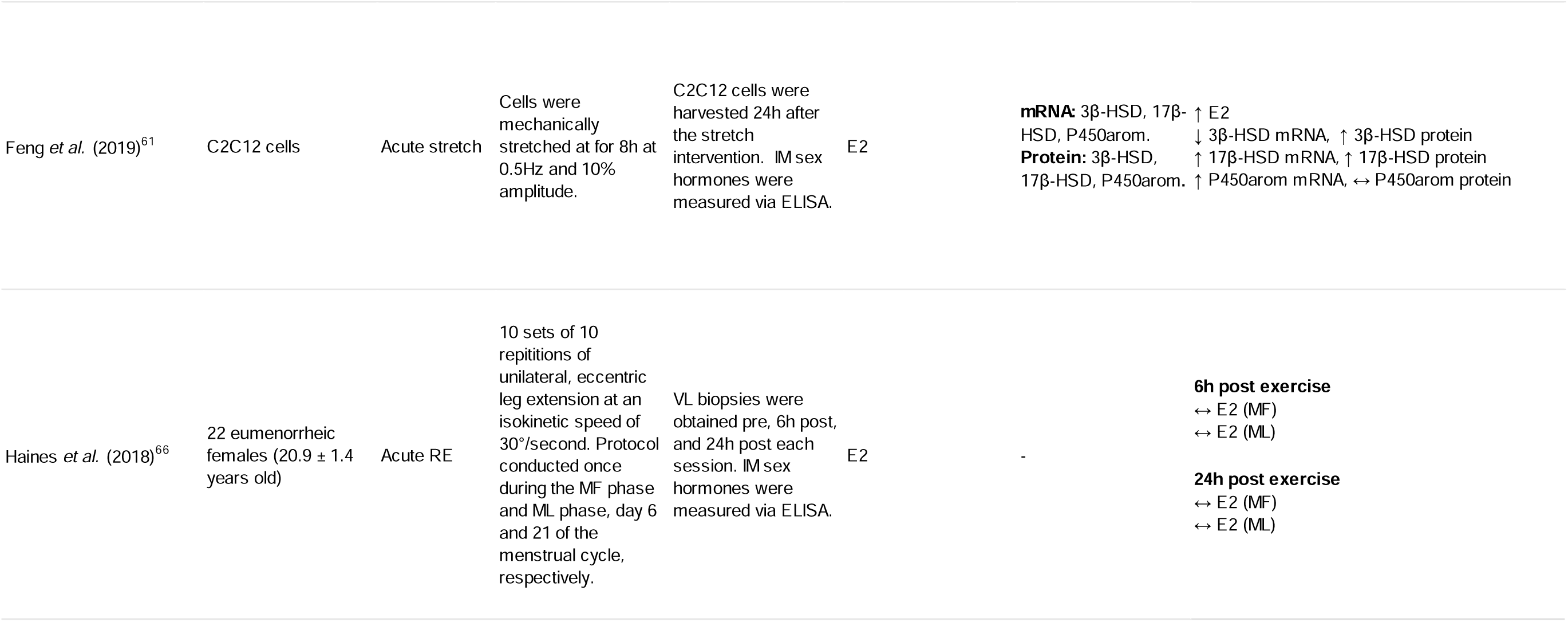
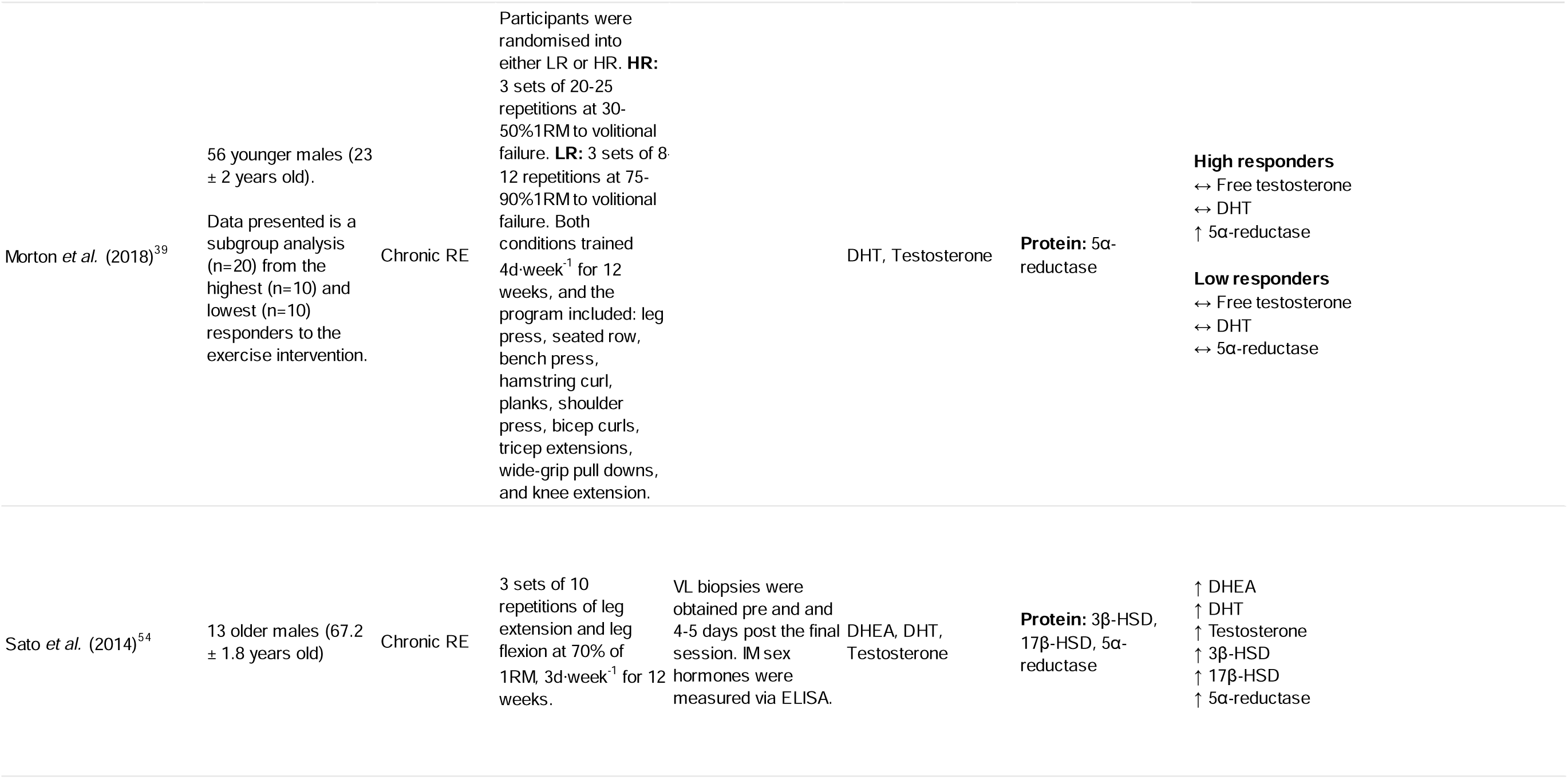
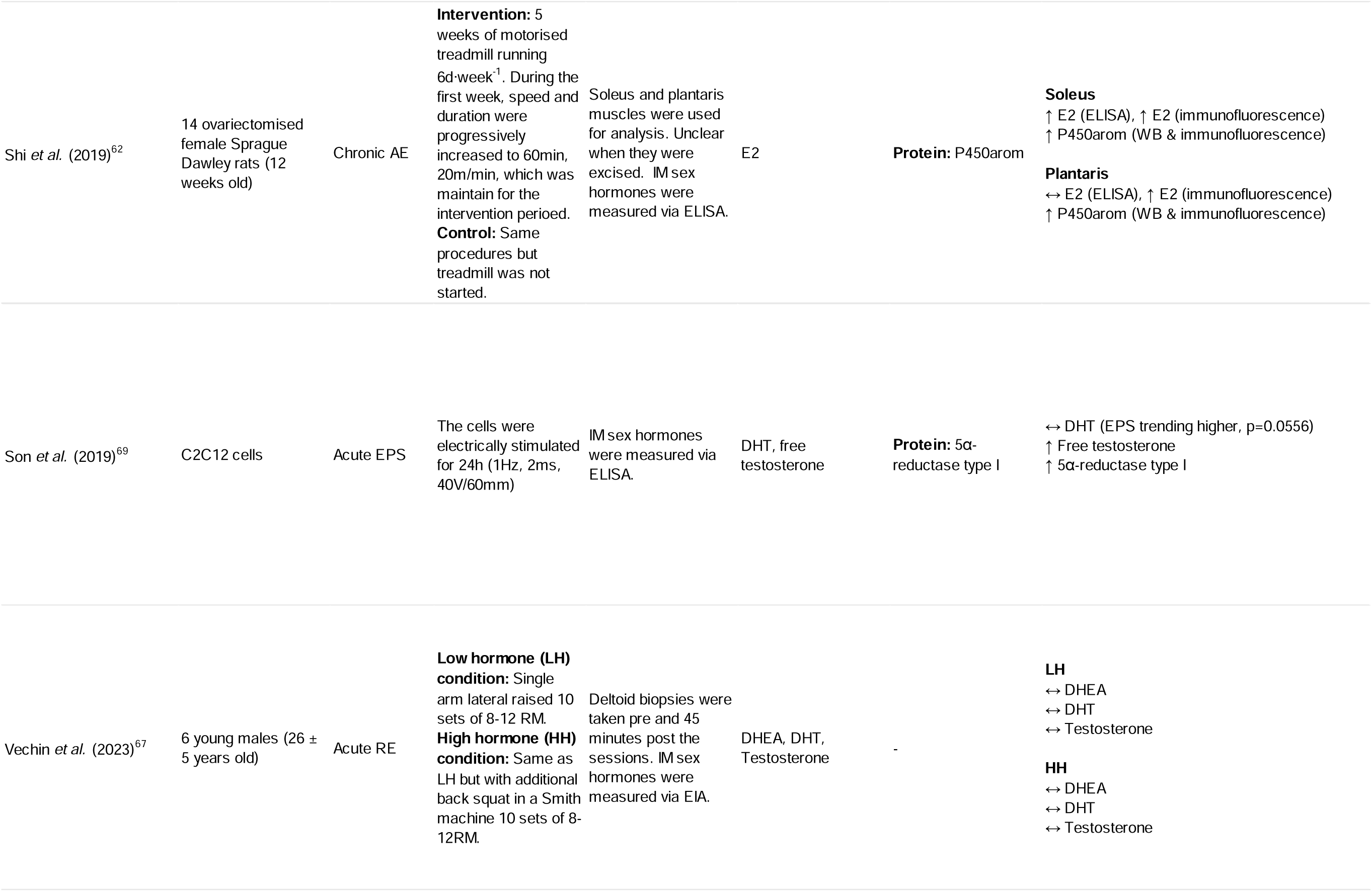
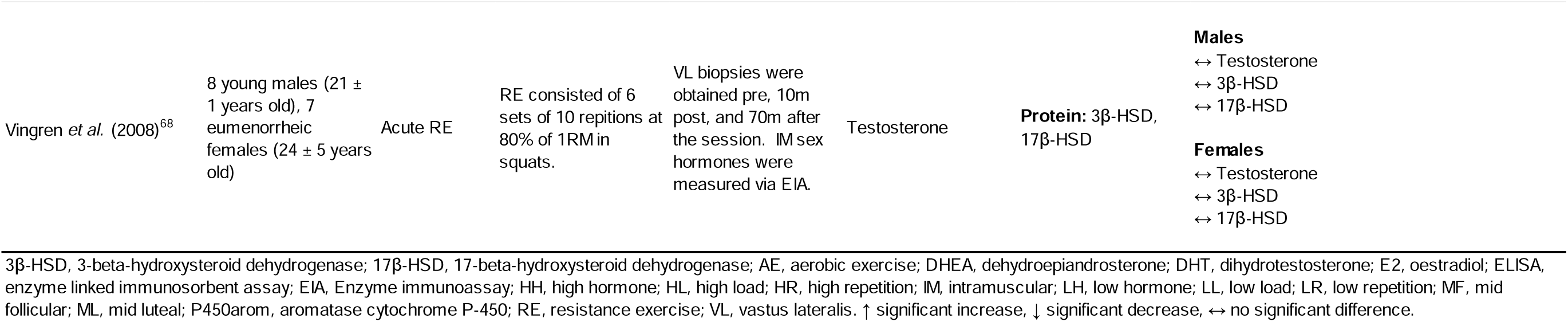
Studies investigating the relationship between exercise and intramuscular sex hormone biosynthesis.

In contrast, in rats, a single session of aerobic training (n=2 studies) increased the intramuscular levels of testosterone, DHT, and DHEA in both male and female rats.^63,64^ Intramuscular E2 increased in male but not female rats. Intramuscular 5α-reductase mRNA levels remained unchanged in males but increased in female rats, while its protein levels increased in both sexes. Both intramuscular P450arom mRNA and protein levels increased in males, but significantly decreased in female rats. Intramuscular 3β-HSD mRNA increased in males but remained unchanged in female rats. Intramuscular 17β-HSD mRNA was unchanged in males but increased in female rats, while intramuscular 17β-HSD protein increased in both.^63,64^

Using a C2C12 myotube model of acute mechanical stretch, Feng *et al.*^61^ found that intracellular E2 levels increased following the intervention, intracellular 3β-HSD mRNA decreased but intracellular 3β-HSD protein increased, intracellular 17β-HSD mRNA and protein increased, and intracellular P450arom mRNA increased but not intracellular P450arom protein. Also in C2C12 cells, Son *et al.*^69^ applied electric pulse stimulation for 24h, which resulted in significant increases in intracellular free testosterone and 5α-reductase protein levels, but not in DHT levels.

#### 4.5.2 Chronic exercise

In a chronic resistance exercise intervention (n=49), subgroup analysis of the ten highest responders (as determined by myofiber CSA and LBM accretion) revealed no effect on intramuscular levels of free testosterone and DHT, but an increase in the 5α-reductase protein levels.^39^ In the ten lowest responders, intramuscular levels of free testosterone, DHT and 5α-reductase protein levels were unaltered.^39^ In older males, chronic resistance training increased the intramuscular levels of testosterone, DHT, and DHEA, as well as intramuscular 3β-HSD, 17β-HSD, and 5α-reductase protein levels following chronic resistance training.^54^ In postmenopausal females, chronic resistance training led to significant decreases in intramuscular DHEA, testosterone, and E2.^45^

In female rats, chronic aerobic training did not affect intramuscular E2 levels in the plantaris muscle but increased in the soleus when analysed using ELISA, whilst intramuscular E2 levels increased in both muscles when analysed using immunofluorescent staining.^62^ Intramuscular P450arom was elevated in both muscles from female rats following exercise.^62^ In male rats, intramuscular total and free testosterone, and DHEA remained unaltered whilst DHT increased after the aerobic training period.^60^ Further, intramuscular 3β-HSD mRNA, and 5α-reductase mRNA and protein and intramuscular P450arom mRNA and protein all increased whilst 17β-HSD mRNA and protein remained unaltered.^60^

## 5 Discussion

In the last two decades, more research has emerged investigating the role and regulation of intramuscular sex hormones, particularly testosterone and E2. Here we aimed to investigate 1) whether intramuscular sex hormones levels vary across the life span in humans, 2) whether intramuscular sex hormones levels are associated with muscle mass and function, or increments in these, and 3) whether exercise impacts intramuscular sex hormone levels and biosynthesis machinery.

### 5.1 Intramuscular sex hormone regulation

It is currently unclear whether there is a relationship between the levels of serum and intramuscular sex hormones, as determination of the biosynthesis location of the sex hormones measured within the muscle remains challenging. However, based on cell, animal, and human research intramuscular sex hormone regulation appears to be decoupled from that of serum hormones and possibly affected by some other signalling factors in serum. Using cultured rat myotubes Aizawa *et al.* (2007)^51^ found that when the cells were incubated with DHEA, a bloodborne testosterone precursor, they synthesised testosterone and E2 in a DHEA dose-dependent manner. This suggests that skeletal muscle has the ability to synthesise these sex hormones in the presence of their precursors. These findings have subsequently been replicated by Pöllänen *et al.* (2015)^59^ in human primary muscle cells. Incubation of cells in pre- or postmenopausal sera resulted in distinct expression patterns of the ERs. However, the ER gene expressions did not differ between cells exposed to the postmenopausal sera from females with elevated E2 due to hormone replacement therapy (HRT) and those without. This suggests that other factors in the sera that varies with menopausal status, other than E2 itself, may affect intracellular hormone signalling.^59^ In line with this hypothesis, they also reported that the intracellular hormone receptors were unaffected by incubation with pure E2.^59^ Adding further complexity, differentiation of myoblasts to myotubes led to increased gene expression of P450arom, ERs and AR, but did not affect E2 levels, indicating that merely increasing the steroid hormone biosynthetic machinery does not lead to increased synthesis of E2.^59^ Hence, the interplay between systemic and intramuscular hormone levels, and the gene expression along their synthesis pathway and their target receptors remains elusive.

In animal models, ovariectomy, that is, the removal of the ovaries to mimic a postmenopausal state, reduces the circulating levels of sex hormones in rats.^70^ Shi *et al.*^62^ reported that 5 weeks of treadmill running in ovariectomised rats increased intramuscular E2 levels. As they did not report circulating E2 levels, it is not entirely clear where these increments originate from. As reported by Zhao *et al.,*^70^ plasma E2 increments occur in the months following ovariectomy, which could potentially increase the intramuscular E2 levels, questioning the validity of ovariectomy as a model of the postmenopausal state. Exercise also promoted the expression of P450arom protein concomitant to the E2 increase, which suggests that the intramuscular increase in E2 might occur from intramuscular origins.

In humans, Pöllänen *et al.* (2015)^59^ found no difference in intramuscular E2 between postmenopausal female twins where one twin used oestrogen containing HRT, and the other one did not. Likewise, following a 12-week progressive resistance training intervention in a group of postmenopausal females receiving either oestrogen treatment or placebo, Dalgaard *et al.*^45^ reported that whilst the serum E2 increased in the oestrogen treatment group, the intramuscular E2 and testosterone remained the same between the treatment and placebo condition. Further, whilst the resistance exercise intervention did not affect the circulating E2 and testosterone levels, the intramuscular E2 and testosterone levels were decreased. Altogether this suggests that intramuscular hormone levels are decoupled from serum hormone levels.

### 5.2 Intramuscular sex hormones and ageing

It is well established that serum E2 concentrations abruptly drop at menopause,^31^ a pattern not conclusively reflected in the muscle. Pöllänen *et al.* (2011)^53^ reported elevated intramuscular E2 levels in postmenopausal females compared to premenopausal counterparts, a pattern however not replicated in another study by the same group,^59^ where intramuscular E2 levels remained unchanged across menopause. Similar inconclusive results were reported for testosterone and DHT in the two studies.^53,59^ Nevertheless, both studies highlighted an increased ratio of intramuscular-to-serum hormone levels in the postmenopausal females compared to their premenopausal counterparts.^53,59^ In contrast, when comparing young and old males, Sato *et al.*^54^ found that intramuscular testosterone, DHT, and DHEA levels mirrored the age-related reduction in the serum hormones.

The reasons for the divergent findings of Pöllänen *et al.* (2011)^53^ and (2015)^59^, and Sato *et al.*^54^ are unclear but there are several differences between the studies that may hinder a robust comparison. The age gap between the younger and older group was substantially greater in Sato *et al.*^54^ than in both Pöllänen *et al.* (2011)^53^ and (2015)^59^. The former cohort also presented significant differences in body composition between the younger and older groups, whilst the pre- and postmenopausal females were anthropometrically matched in both Pöllänen *et al.* (2011) and (2015).^53,54,59^ Both age and body composition are well established moderators of systemic sex hormone levels.^18,71^ Although intramuscular sex hormone levels appear to be regulated somewhat independently from the systemic sex hormones, one cannot exclude the possibility that age- and body composition affect the intramuscular sex hormones too, perhaps in a sex-specific manner. Future studies should control for these putative confounding variables.

### 5.3 Intramuscular sex hormones and their association with muscle mass and function

Associations between circulating physiological levels of testosterone and lean body mass have been found in males,^22^ but not females,^24^ with some equivocal findings in the relationship between E2 and muscle mass in females.^24^ In postmenopausal females, the relationship between intramuscular sex hormones and muscle mass reflected the circulating hormones, as intramuscular testosterone and E2 were not associated with muscle mass.^24,32,45^ However, similar to circulating androgens, intramuscular androgens were associated with muscle CSA, but only in older males.^22,39,54^

Following an exercise intervention, it was found that exercise induced increments in intramuscular androgens and E2 were correlated to muscle mass and muscle cross sectional area in older males, and male and female rats,^54,60,62^ but not younger males or postmenopausal females.^39,45^ In females, increased intramuscular sex hormone levels were associated with a greater proportion of fat infiltration.^53^ Notably, the aforementioned human trials found that the higher intramuscular testosterone levels the lower the muscle mass and the greater the loss of muscle quality (increased fat infiltration), while circulating testosterone levels did not correlate with either muscle mass or muscle quality.^53,54^ Fat tissue has a steroidogenic capacity, which appears to increase with age.^72^ Likewise, intramuscular fat infiltration also increases with age,^73^ making it possible that the associations between greater fat infiltration and intramuscular sex hormones originate from increased hormonal biosynthesis from the fat tissue. However, in a pooled sample of postmenopausal females randomised to either transdermal oestrogen replacement therapy or placebo and undergoing a resistance training intervention, intramuscular E2 and testosterone levels significantly decreased despite unaltered intramuscular fat levels. Further, no association between intramuscular sex hormones and intramuscular fat content was found,^45^ altogether refuting the notion that the fat infiltration is the main determinant of intramuscular sex hormone levels.

Absolute knee extensor strength showed significant positive associations with several androgens in males and females, and E2 in females.^54,59^ In females, these associations disappeared when normalised to muscle CSA.^53^ However, even though Dalgaard *et al.*^45^ found that both intramuscular testosterone and E2 decreased following a 12-week resistance training intervention in postmenopausal females, they did find that the better the maintenance of these intramuscular hormones, the greater the improvement of muscle function. Whilst Pöllänen *et al.* (2015)^59^ reported their strongest associations between intramuscular sex hormones and vertical jump height in females, suggesting that the intramuscular hormones may modulate muscle contractile properties. Dalgaard *et al.*^45^ found a numerical but non-significant association between intramuscular testosterone and E2, where the maintenance of testosterone and E2 were positively correlated with increased vertical jump height, albeit with a smaller cohort increasing the risk for a type II error. Generally, animal research supports the plausible association between muscle contractile properties and intramuscular hormones, where avian and rodent research both indicate that fast twitch muscle phenotypes tend to express higher levels of the intramuscular sex hormones and the associated biosynthetic enzymes than more slow twitch muscle phenotypes.^74^ Currently, intramuscular sex hormones’ putative effect on contractile properties remains speculative. Future research should consider human fibre type specific measures of intramuscular sex hormones.

### 5.4 Exercise to manipulate intramuscular sex hormone levels

A single resistance exercise training session did not affect intramuscular sex hormone levels in human, with all five studies included reporting no effect on intramuscular hormone levels and the gene and/or protein expression of relevant enzymes along the synthesis pathway.^45,65–68^ In adult males, the most pronounced post-exercise elevation in circulating total testosterone is seen immediately following exercise cessation with a decrease of 0.015 nmol/L for each minute that follows thereafter.^75^ The included trials conducted post exercise biopsies from 10 minutes to 24 hours after the final exercise session, with only one study measuring the intramuscular hormone levels within 30 minutes following exercise, which is when the transient increase in serum testosterone typically returns to baseline.^75^ Contrasting the results of the human trials, single bouts of running exercise were a potent stimulus to increase intramuscular androgen levels in both male and female rats, and E2 in male but not female rats.^63,64^ Compared to the acute human trials, the exercising rats’ muscles was excised immediately following exercise cessation. Whether temporal discrepancies in post-exercise muscle collection timing between the human and animal studies may explain the divergent findings remain unknown. The interventions also differed between humans and rats, where the latter were all exposed to aerobic exercise, which tends to yield a different molecular signalling pattern than resistance training.^76^ Considering the timeframe of the transient post-exercise elevation in circulating testosterone and the immediate increments in intramuscular sex hormones in rats, future research should consider collecting human biopsies directly following intense exercise.

In contrast to acute exercise interventions, 12 weeks of progressive resistance training led to reductions in intramuscular testosterone, DHEA, and E2 following the intervention in postmenopausal females.^45^ In elderly males, chronic resistance training increased intramuscular levels of testosterone, DHT, and DHEA following a similar intervention,^54^ whilst androgens remained unchanged in younger males following a resistance training intervention.^39^ Given that Sato *et al.*^54^ found that ageing led to a reduction in intramuscular sex hormones, and that exercise increased the intramuscular hormone levels in older participants towards levels seen in the younger participants, one could speculate that the lack of effect of resistance exercise on intramuscular sex hormones reported in Morton *et al.*^39^ may be as their younger participants already had normal hormone levels, with exercise serving only to restore geriatric losses.

Both chronic exercise studies in rodents used aerobic treadmill running as their interventions.^60,62^ Consequently, aerobic exercise caused intramuscular elevations in E2 in the soleus muscle in ovariectomised rats, but not in the plantaris from the same rats nor the gastrocnemius from male rats. Further, whilst DHT increased, DHEA and testosterone remained unchanged in the trained male rats compared to their sedentary counterparts. Studies are yet to measure intramuscular androgens in female rodent models following chronic exercise interventions.

### 5.5 Methodological considerations

At present, the methodology used to measure intramuscular sex hormones may not be sufficiently sensitive for accurate measures, particularly at low hormonal concentrations. Most studies used ELISA/EIA, which can both under or overestimate hormone levels, particularly at low concentrations.^77,78^ Compared to immunoassays, liquid chromatography mass spectrometry (LC/MS) yields a significantly lower variability and minimises measurement error in biological fluids.^77^ Whether this measurement error at relatively low concentrations is reflected in the muscle remains untested. One can however surmise that it would be similar, as the same pattern is reflected in other matrices such as urine and saliva.^79,80^

To the best of the authors’ knowledge, no method has yet been validated to measure intramuscular sex hormones. Hence, there is a need for standardised and validated methodology to ensure valid and reliable measures of the intramuscular sex hormones and facilitate the comparison of findings between studies. Such methodology will likely be based on mass spectrometry, the gold standard of hormonal measurements in other matrices.^81^

### 5.6 Conclusions

Whilst the research field of intramuscular sex hormones present mostly inconclusive and contradictory findings between studies, the possible relationships between skeletal muscle mass and function and intramuscular hormones warrant further investigation, with considerations into age and sex as potential confounding variables.

## Data Availability

All data produced in the present study are available upon reasonable request to the authors

## 6 Acknowledgements

Séverine Lamon is supported by an Australian Research Council Future Fellowship (FT210100278).

