## Supplementary material for "The Role and Regulation of Intramuscular Sex Hormones in Skeletal Muscle: A Systematic Review"

**Supplementary table S1:** Search strategy for systematic review

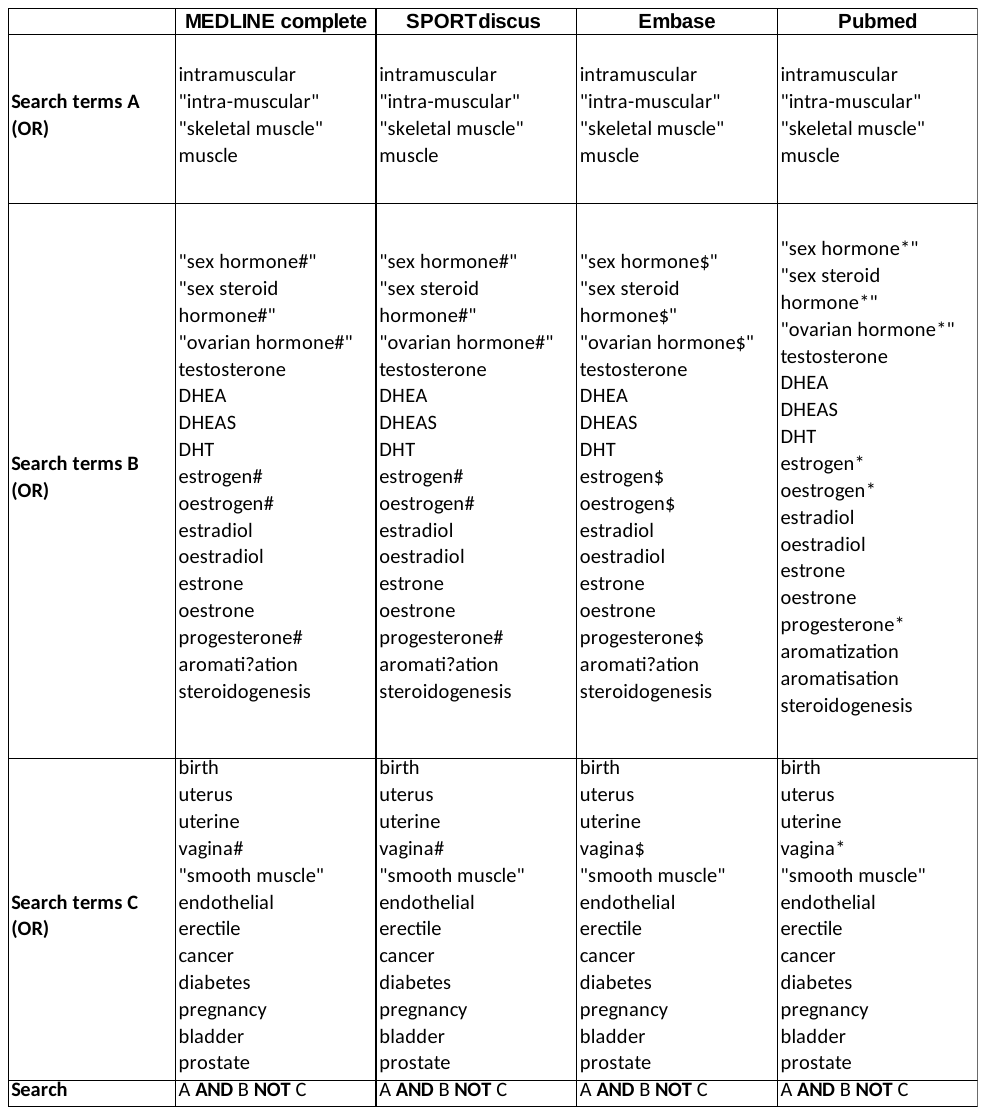

**Supplementary table S2:** PICO model for systematic review eligibility

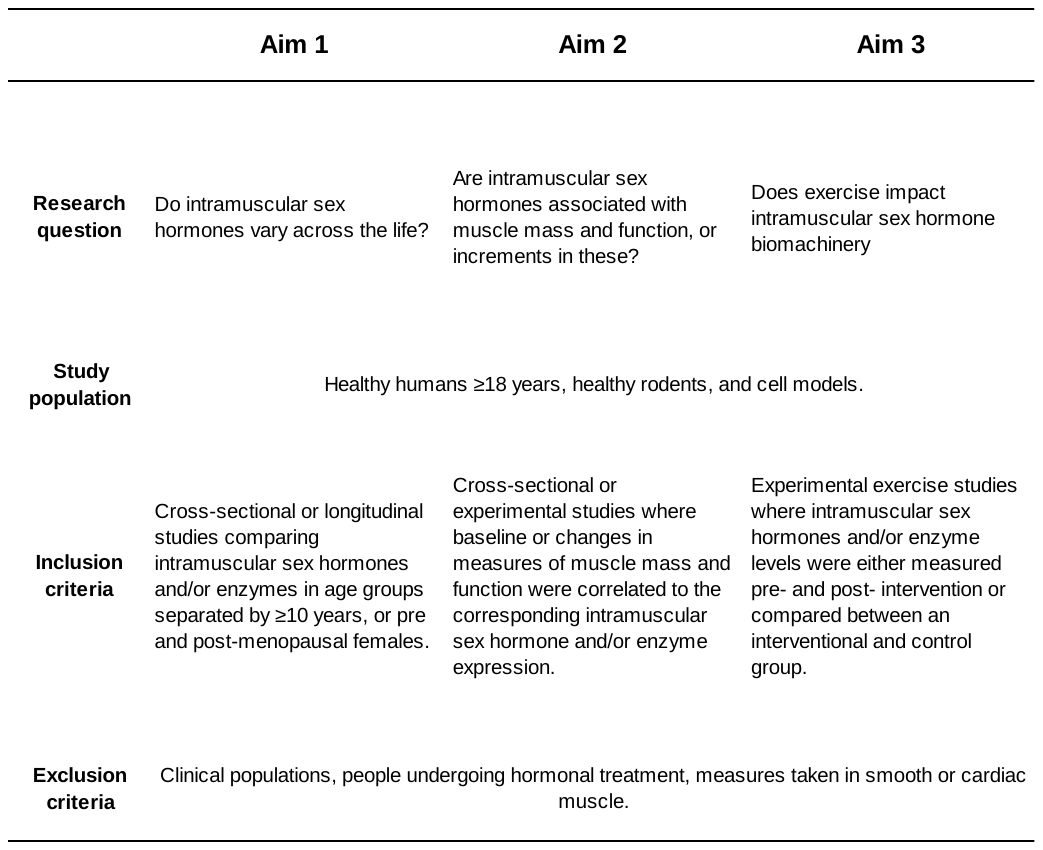

**S3. Quality assessment of cross-sectional studies**

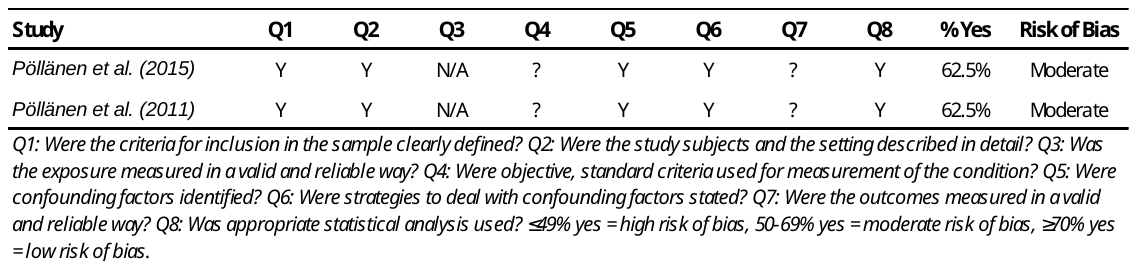

**S4. Quality assessment of randomised controlled trials**

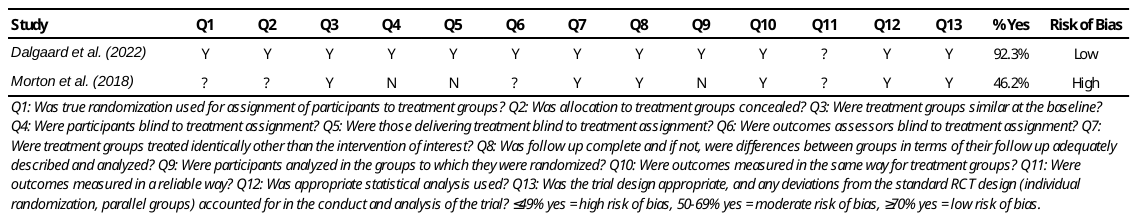

**S5. Quality assessment of quasi experimental studies**

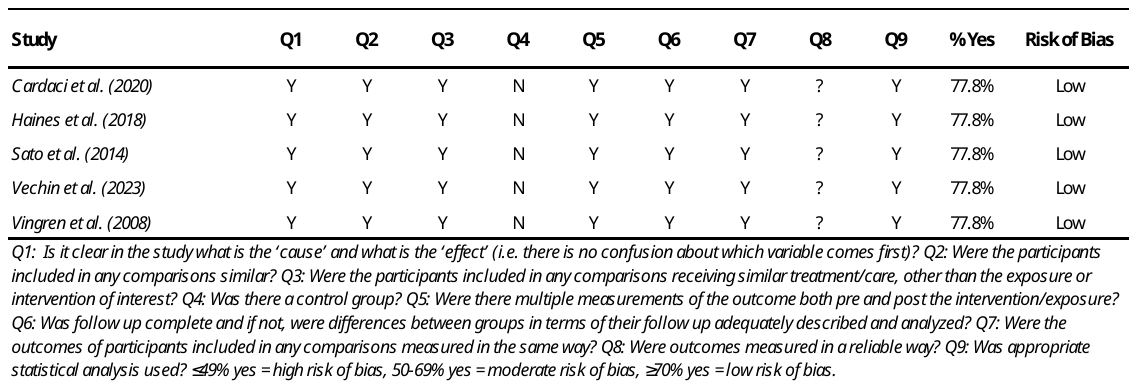

**S6. Quality assessment of animal trials**

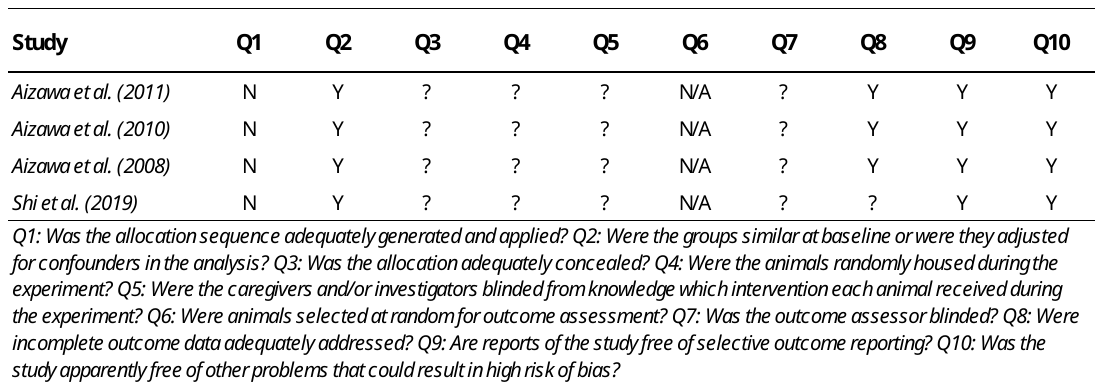

**S7. Quality assessment of *in vitro* trials**

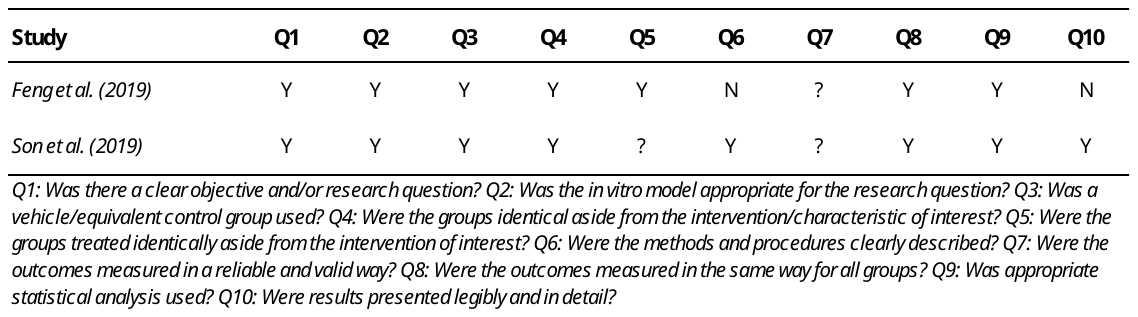
